# Factors influencing the delayed presentation of patients with large vessel occlusion in acute ischemic stroke

**DOI:** 10.1101/2023.10.27.23297702

**Authors:** Gan-Ji Hong, Shao-Hui Zeng, Ting-Yu Yi, Yan-Min Wu, Ding-Lai Lin, Zhi-Nan Pan, Xiu-Fen Zheng, Xiao-Hui Lin, Rong-Cheng Chen, Li-San Zeng, Jin-hua Ye, Wen-Huo Chen

## Abstract

**Objective:** The benefit of treatment for patients with large vessel occlusion (LVO) in acute ischemic stroke (AIS) is associated with timely access to reperfusion therapy. This study aimed to analyze the factors associated with delayed access to medical attention among patients with AIS-LVO.

**Methods:** Patients with acute ischemic stroke admitted to the cerebrovascular intervention department of Zhangzhou Hospital, Fujian Province, China, from September 1, 2020 to September 1, 2022, were enrolled in this study. Patients with AIS-LVO were divided into two groups based on whether they visited the hospital for more than 24 hours. Demographic data, NIHSS scores, clinical and radiological data, and in-hospital flow time were collected. Differences between the two groups were analyzed, and univariate and multivariate analyses were performed to identify factors associated with delayed presentation.

**Results:** A total of 1396 patients were included in the study, with 218 in the delayed group and 1178 in the non-delayed group. There were no significant differences between the two groups in terms of age, gender, or education level. However, the delayed group had a higher proportion of patients living alone (*P*<0.05). Regarding stroke risk factors, hypertension (83.5% vs 69.9%, *P*<0.001), diabetes mellitus (35.8% vs 26.1%, *P*=0.047), and smoking (39.5% vs 28.0%, *P*=0.020) were more prevalent in the delayed group compared with the non-delayed group. Conversely, the non-delayed group had a higher incidence of atrial fibrillation (37.5% vs 12.8%, *P*<0.001). After multivariate regression analysis, the independent predictors for the delay group were solitary OR=11.10 (95% CI 5.72-21.5), large artery atherosclerosis OR=2.63 (95% CI 1.40-4.94), presence of atrial fibrillation OR=0.41 (95% CI 0.19-0.87), and involvement of the middle cerebral artery OR=0.51 (95% CI 0.32-0.81).

**Conclusions:** Among patients with AIS-LVO who delay seeking medical attention for more than 24 hours, large artery atherosclerosis appears to be the predominant pathogenesis.

## INTRODUCTION

The prevalence of large vessel occlusion (LVO) in acute ischemic stroke (AIS) is 13-52%, [1] and it is considered a severe subtype that independently predicts poor outcome at 6 months post-stroke.[2] Endovascular therapy (EVT) has demonstrated effectiveness in achieving early LVO reperfusion,[3-5] but current guidelines, such as DAWN, only recommend EVT for patients within 24 hours of symptom onset.[6] Timely salvage of AIS-LVO is essential for improving patient prognosis.

The process leading up to AIS reperfusion therapy involves both pre-admission and post-admission phases. The pre-hospital phase consists of the patient’s arrival at the hospital following the onset of stroke, either through self-transportation or emergency medical services. The post-hospital phase involves the implementation of the green channel strategy for stroke, which aims to facilitate the rescue process and shorten the in-hospital resuscitation time. Delays in either the pre-hospital or in-hospital phases can result in patients missing the optimal time window for reperfusion therapy.[7]

Previous studies have found that delay in stroke care is associated with living alone and nocturnal stroke episodes.[8] In China, social living conditions and access to health care are the main factors influencing prehospital delay in patients with acute ischemic stroke.[9] However, the potential factors contributing to delays in care for patients with AIS-LVO remain unclear. In this study, we aimed to compare the demographic characteristics of LVO patients, as well as pre-hospital and post-admission flow data within 24 hours of symptom onset and between 24 and 72 hours, in order to explore factors associated with delays in seeking care beyond 24 hours. It is hoped that in the future, overcoming these factors will help to optimize the reperfusion rescue strategy for LVO patients.

## METHODS

Our center is located in the southeast of China, serving a population of approximately 5 million people. It is a comprehensive stroke center equipped with both thrombolytic and EVT capabilities. Our emergency department operates with a 24/7/365 mobile stroke team and a regional referral network for stroke care based on geographic location and resource allocation. Upon arrival at the Emergency Department, patients are promptly assessed and triaged by the emergency nurses. Those suspected of having a stroke after evaluation by neurologists then undergo the stroke greenway assessment process for basic blood tests, electrocardiogram, multimodality imaging exams include cranial computed tomography (CT), computed tomography angiography (CTA) and computed tomography perfusion (CTP). Further management is performed based on the results.

### Inclusion and exclusion criteria

A prospective registry (September 2020 to September 2022) of patients with acute ischaemic stroke through the emergency green channel at Zhangzhou Affiliated Hospital of Fujian Medical University (n=4876) was conducted. Of these patients, 1662 were assessed by multimodal CT imaging for the presence of head and neck macrovascular lesions. Stroke patients with an onset time of more than 72 hours were excluded (n=158), and patients with non-responsible large vessel occlusion or just stenosis were excluded (n=268). Ultimately, 1396 patients met the clinical criteria and were included in the analysis if their onset of stroke symptoms occurred within 72 hours prior to admission, or if the time considered as last normal to the time of admission fell within this timeframe and were identified as having a responsible large vessel occlusion (in the intracranial or extracranial carotid artery, proximal to the middle cerebral artery M1 or M2, vertebrobasilar artery, and proximal to the anterior or posterior cerebral artery) based on multimodal CT imaging. They were divided into delayed (n=218) and non-delayed groups (n=1178) according to whether the onset occurred within the first 24 hours. The inclusion and exclusion criteria are shown in Figure 1.

**Figure 1.**
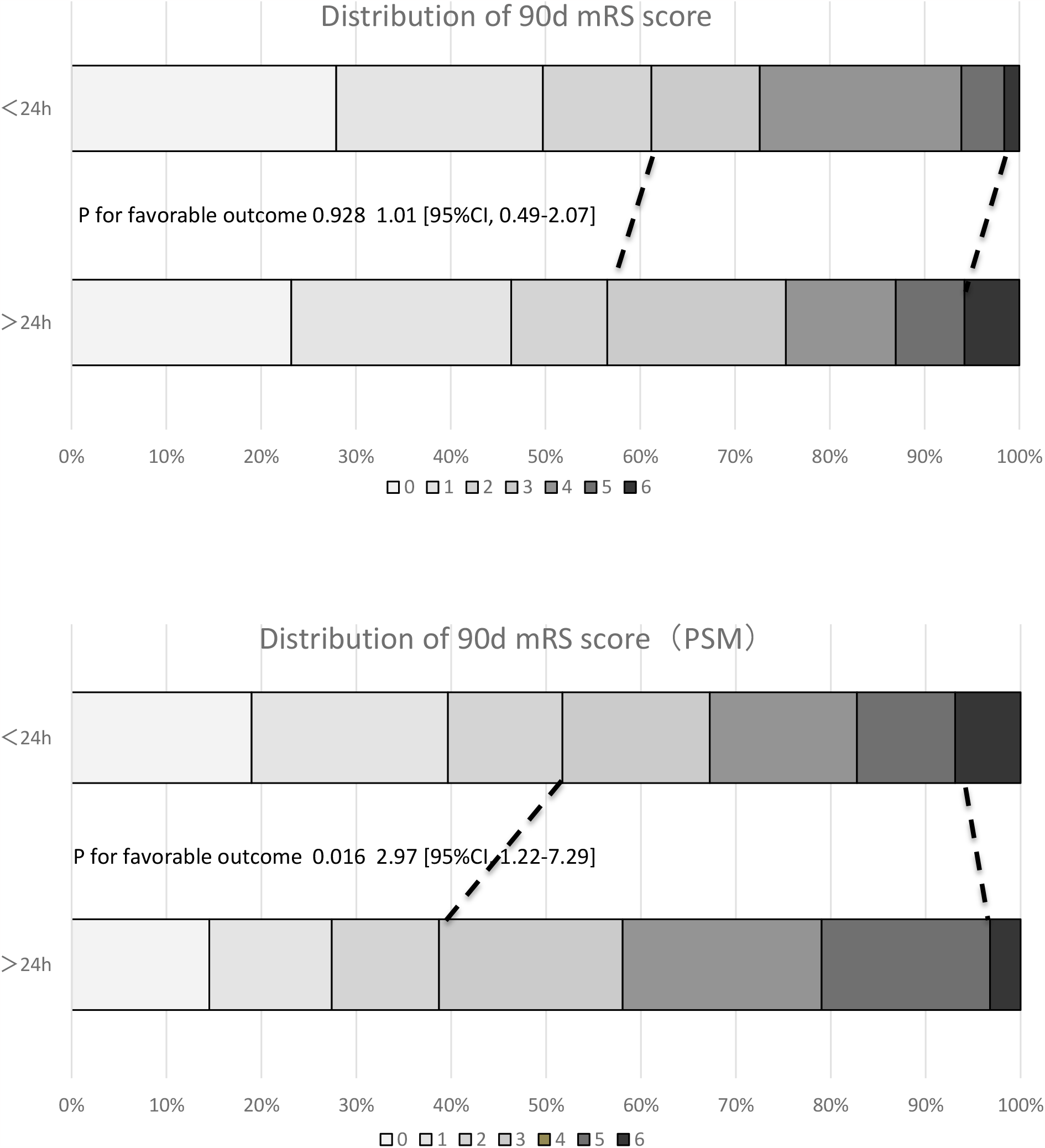

### Data Collection

Demographics data of the patients including gender, age, residence status, education level, and mode of hospital visit were collected. Clinical data, including mRs (modified Rankin Scale score) score before onset, NIHSS score on admission, history of previous cerebral infarction disease, hypertension, diabetes, hyperlipidemia, atrial fibrillation, history of smoking, and whether they experienced progression of the disease were also collected. Mechanism of morbidity (based on TOAST), the mRs at discharge, and NIHSS score at discharge were collected according to the registry medical record. The mRs after 90 days were obtained by outpatient assessment or telephone follow-up. Functional independence was defined as an mRs 0-2 at 90 days.

Obtaining Alberta Stroke Programme Early CT Score (ASPECTS) and responsible occluded vessels were determined based on cranial CT and CTA imaging results. CTP were automatically estimated using perfusion imaging selection criteria provided by the RAPID software (iSchema View) for core infarct volume and ischemic penumbra volume (ischemic tissue volume was the sum of core infarct volume and ischemic penumbra volume). The mismatch ratio was defined as the ratio of ischemic tissue volume to core infarct volume. To eliminate bias, the collected data were assessed by both a neurologist and a neuroradiologist.

Data were collected at various stages of the patient’s admission and EVT procedure workflow, including time to CT examination, time to groin puncture, time to first reperfusion, and procedure length. The time to first reperfusion was defined as the time when the angiography suggested the initial recanalization of blood flow in the target vessel.

### Statistical Analysis

Nominal variables are described as frequencies, while continuous variables are reported as medians and interquartile ranges (IQR). Categorical variables are presented as frequencies and percentages. Pearson chi-square tests or Fisher exact tests for categorical variables were used to compare differences in baseline characteristics between the above categories. Multivariate logistic regression analysis was performed to assess independent risk factors in patients with large vessel occlusion beyond 24 hours of onset. To eliminate possible confounding factors between the delayed and non-delayed groups, a 1:1 matching was performed using propensity score analysis to compare the two functionally independent results. All *P* values were bilateral, and *P* < 0.05 was considered statistically significant. All statistical analyses were performed using SPSS 27.0.1 software.

## RESULTS

A total of 1396 patients were included in the study, 218 in the delayed group and 1178 in the non-delayed group. There were no significant differences between the two groups in terms of age, gender, or education level, and the delayed group had more patients living alone (3.9% vs 29.4%, *P*<0.001). There was no significant difference between the two groups in terms of the mode of consultation and whether they were transported by pre-hospital emergency.

Regarding stroke risk factors, hypertension (69.9% vs 83.5%, *P* <0.001), diabetes mellitus (26.1% vs 35.8%, *P* =0.047), and smoking (28.0% vs 39.5%, *P* =0.020) were more common in the delayed group compared with the non-delayed group. The non-delayed group had a higher incidence ofatrial fibrillation (37.5% vs 12.8%, *P* <0.001). The delayed group had a greater proportion of large-artery atherosclerotic cerebral infarcts in the pathogenesis of stroke (46.0% vs 82.6%, *P* < 0.001). There was no significant difference between the two groups in pre-onset mRs scores, but the non-delayed group had higher NIHSS scores at admission (median 14 vs. 8 *P* < 0.001). The delayed group experienced more progressive exacerbation of symptoms (6.8% vs 28.0%, *P* = 0.031). Endovascular treatment was administered to 130 patients (59.6%) in the delayed group and 1026 patients (87.1%) in the non-delayed group, with a significant difference between the two groups.

Regarding imaging findings, the admission-based cranial CT plain scan ASPECTS score was higher in the delayed group than in the non-delayed group (7 vs 8, *P* = 0.017). In terms of involved vessels, occlusion of the middle cerebral artery was more common in the non-delayed group (55.0% vs 36.7%, *P* = 0.011), with no significant difference between the two groups for the remaining vessel involvement. The non-delayed group had a larger core infarct volume (median 16 vs 3, *P* = 0.014) and a larger ischemic penumbra (median 81 vs 62, *P* = 0.023) but a smaller mismatch ratio (median 5.4% vs 11.2%, *P* = 0.040) than the delayed group (Table 1). At the time point of the in-hospital procedure, the two groups did not reach statistical significance in terms of arrival to CT examination time, arrival to puncture time, arrival to reperfusion time, and procedure length (Table 2).

**Table 1.**
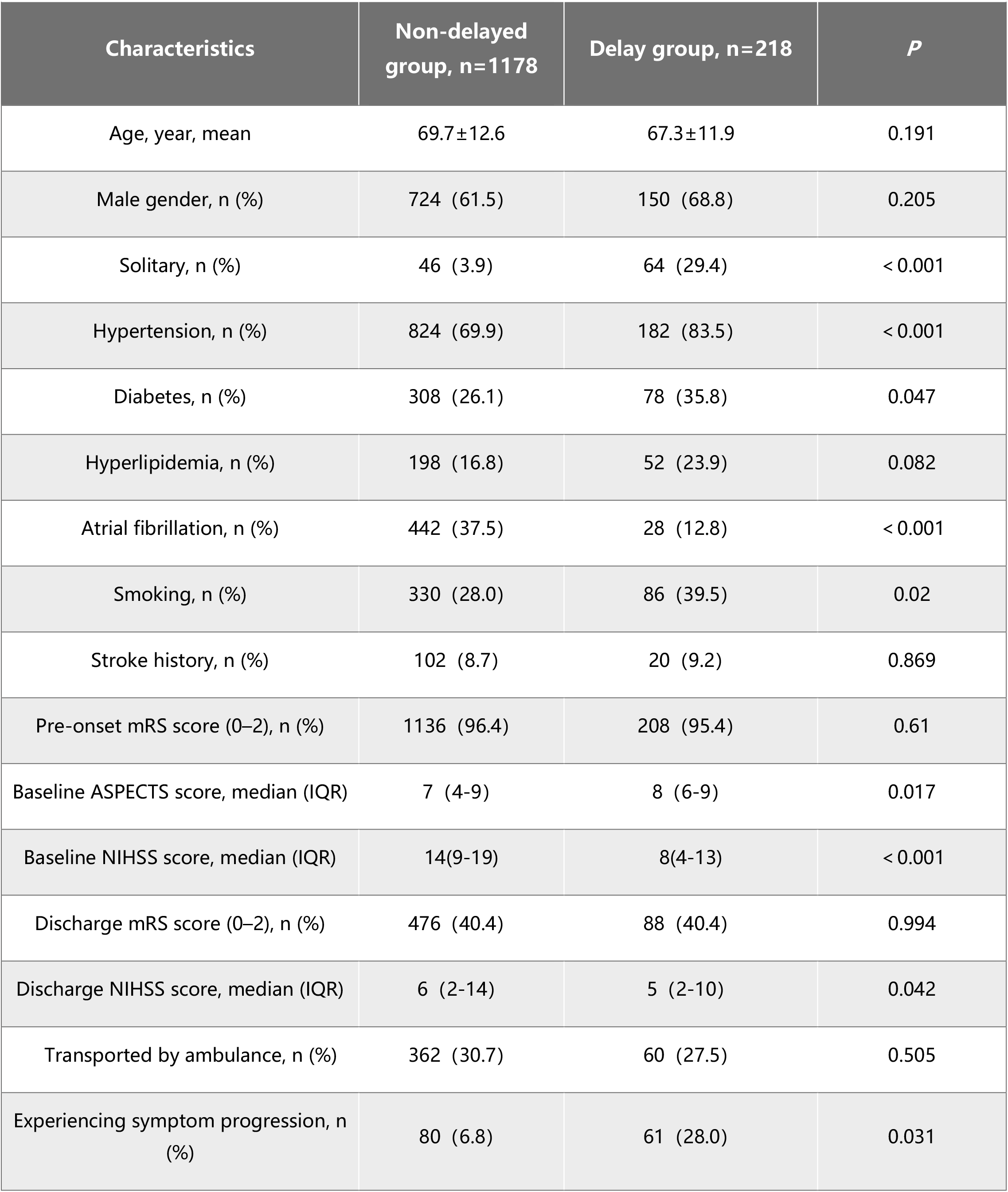
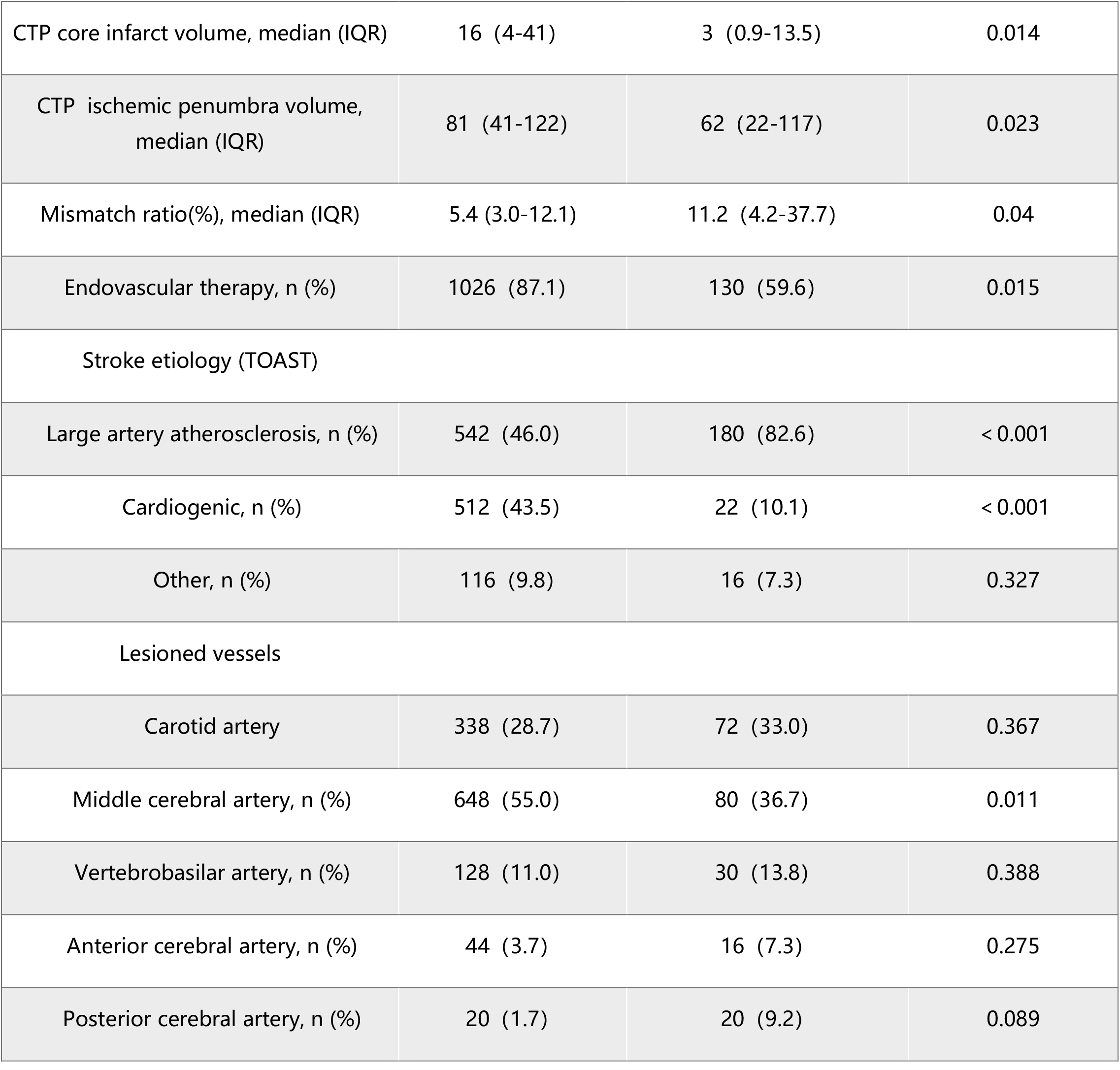

**Table 2.**

| Time course | Non-delayed group, n=1178 | Delay group, n=218 | *P* |
| --- | --- | --- | --- |
| Onset-to-door time, min, median (IQR) | 363 (195.0-706.5) | 2196 (1681.5-2979.0) | < 0.001 |
| Door-to-CT time, min, median (IQR) | 20 (15-27) | 20.5 (14-33.5) | 0.387 |
| Door-to-puncture, min, median (IQR) | 96 (81.0-118.8) | 110 (90.0-148.3) | 0.381 |
| Door-to-perfusion, min, median (IQR) | 149 (123.0-199.3) | 188 (164-232) | 0.138 |
| Duration of surgery, min, median (IQR) | 99 (31-374) | 112 (75-137) | 0.270 |

Multivariate regression analysis identified independent predictors for the delayed group, including living alone OR=11.10 (95% CI 5.72-21.5), large artery atherosclerosis OR=2.63 (95% CI 1.40-4.94), presence of atrial fibrillation OR=0.41 (95% CI 0.19-0.87), involvement of middle cerebral artery OR=0.51 (95% CI 0.32-0.81) (Table 3). After propensity score matching, 124 cases were in the delayed group and 124 in the non-delayed group, and all baseline covariates were statistically indistinguishable except for time to visit. After adjustment, the non-delayed group was more functionally independent at 90 days and statistically different between the two groups (Figure 2).

**Table 3.**
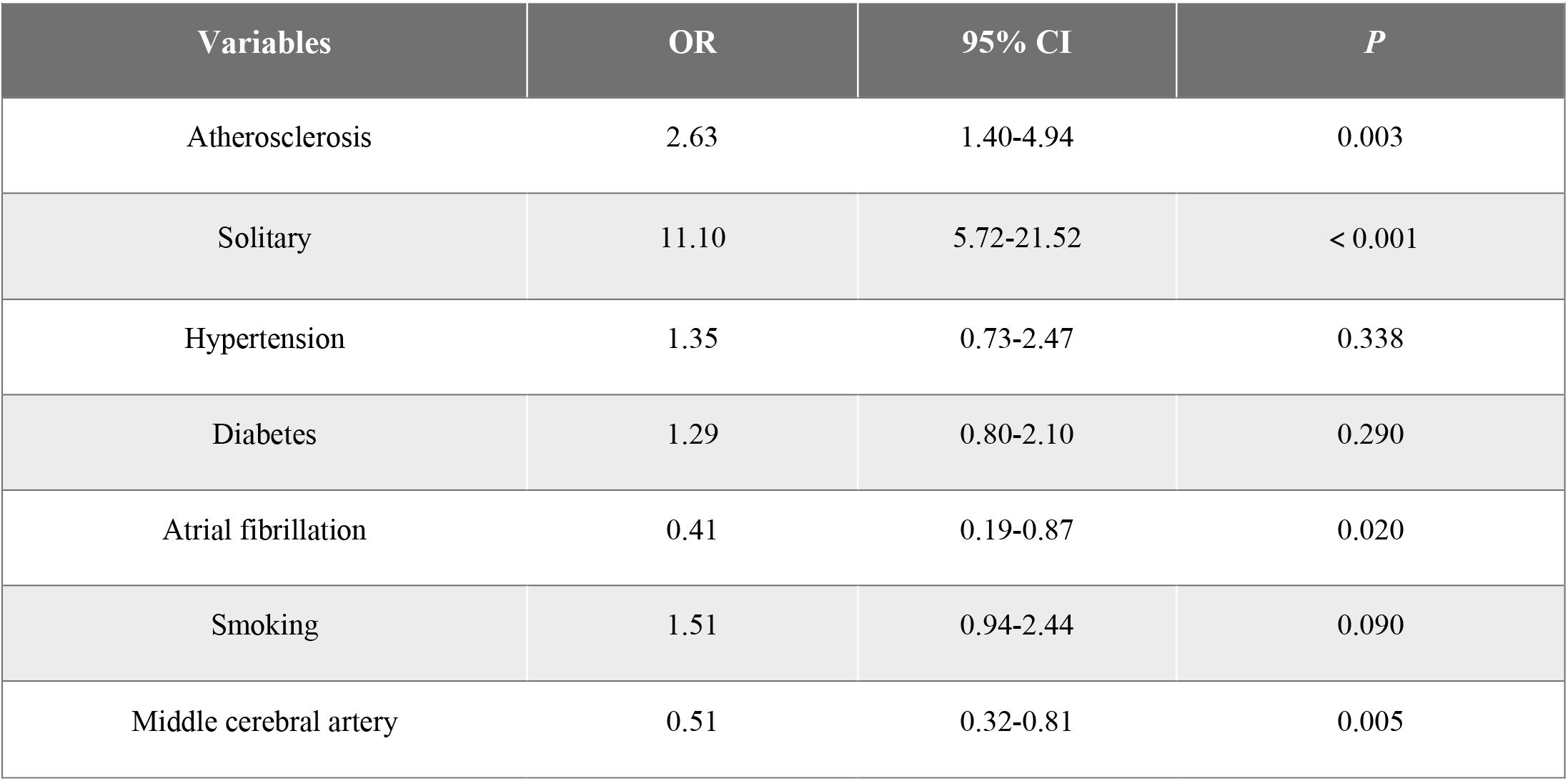

## DISCUSSION

We are, to our knowledge, the first study to explore the characteristics of the AIS-LVO population with delayed attendance, and the results suggest that patient treatment delays are characterized by large artery atherosclerosis, live alone, and have relatively mild early symptoms.During the COVID-19 epidemic, delays in stroke treatment have been observed and the treatment of patients with AIS-LVO is affected to varying degrees. The time from onset to consultation was prolonged and DNT for thrombolytic therapy was delayed compared to pre-COVID-19.[10, 11]. As the data were all collected during the pandemic period, we were able to account for the epidemic’s influence on the process. In contrast, studies conducted during this period only addressed factors affecting stroke flow delays.

This study suggests that people with AIS-LVO presenting beyond 24 hours had a higher prevalence of atherosclerosis-related risk factors, such as hypertension and diabetes, and a lower proportion of atrial fibrillation. Our results also support the notion that t the delayed group predominantly consists of individuals with large artery atherosclerosis infarctions. Our results showed that the baseline NIHSS scores were higher in the non-delayed group than in the delayed group, probably due to the higher proportion of cardiogenic embolic cerebral infarctions in the non-delayed group, which generally have severe symptoms at the beginning of the onset of the disease, resulting in patients being seen as soon as possible after the onset of the disease[12]. However, the results suggest that the delayed group had lower admission ASPECT scores, this finding does not imply that infarctions were more severe in the delayed group than in the non-delayed group, as hypointense lesions are less visible on CT during the hyperacute phase. In terms of vascular involvement, the delayed group had a higher prevalence of the involvement of the middle cerebral artery, and previous studies have found a higher proportion of large artery atherosclerosis in the pathogenesis of middle cerebral artery infarction[13], which may also support a higher proportion of large artery atherosclerosis types in the delayed group.

A study from the USA, analyzing multicenter data from 2002 to 2021, found that almost 40% of cases arrived at the hospital within 3 hours of onset, with an overall median time from onset to presentation of 4.7 (interquartile range, 1.5-12.7) hours.[14] Only 84.4% of patients with AIS-LVO in our center visited a hospital within 24 hours of onset. In contrast, in an interview study involving patients within 72 hours of stroke onset, 38% said they were aware of the warning signs of stroke, but only 25% correctly identified their symptoms as stroke symptoms[15]. Our findings show that 27.5% of patients experienced progression of symptoms early in the onset of stroke before they chose to visit a hospital or were referred to a more advanced hospital. The results of the study showed that patients in the delayed group had smaller infarct core volumes, larger ischemic semi-dark zone volumes and mismatch ratios, explaining the tendency for the onset of symptoms to be mild, leading patients or family members to downplay stroke symptoms, and that patients who acknowledge the severity of symptoms have shorter pre-hospital delays than those who do not. In China, public health education about stroke is still inadequate and many people are still not sufficiently aware of the dangers of stroke.

Previous studies have shown that living alone is a risk factor for pre-hospital delay at the time of first presentation of stroke symptoms.[16] The present study suggests that living alone is an independent risk factor for delayed treatment of AIS-LVO. Bystanders recognize the symptoms of stroke more often than the patients, who themselves more often use a wait-and-see approach to seeking treatment.[17] And as most of the people living alone in our region are elderly or unmarried, infertile middle-aged men, society should pay more attention to this group.

Previous retrospective cohort studies have suggested that earlier access to treatment for AIS is associated with improved outcomes. Each 15-minute reduction in treatment time has been linked to higher rates of independent discharge, better functional independence upon discharge, and reduced mortality [18]. Through propensity-matched analysis, we found that the delayed group had a poorer 90-day prognosis compared to the non-delayed group, emphasizing the importance of early treatmen to patients with cerebral infarction for better outcome and prognosis. The DIFFUSE3 trial confirmed that acute ischemic stroke patients with a core infarct volume of <70 ml, an ischemic semi-dark zone volume of >15 ml and a mismatch ratio of >1.8 still had a benefit from 6-16 h endovascular treatment[19]. A proportion of AIS-LVO patients at our center with an onset beyond 24 hours still have small infarct cores and high mismatch ratios, and this time-delayed population may still benefit from EVT after rigorous screening with imaging. However, this needs to be validated in a larger sample and in a multicenter prospective randomized study.

There are some limitations to our study, firstly this was a single-center study conducted in China, where atherosclerosis is inherently more common [20] and the conclusions may not be generalizable to other national regions. Secondly, the lack of specific analysis of the impact of different treatment modalities on prognosis in the study may have potentially biased the results. Thirdly, the delayed group does not distinguish in detail the causes of delays that are due to delays in referrals from other hospitals, although the proportion of patients in this category is small in our center which may have contributed to the bias in the results.

## CONCLUSION

In our center, among patients with AIS-LVO who delay seeking medical attention for more than 24 hours, large artery atherosclerosis appears to be the predominant pathogenesis. they tend to live alone and have relatively mild symptoms in the early stages, often experience progression before choosing to present to hospital. Public education on stroke must be strengthened in the future, especially for those at high risk of delayed treatment.

## Data Availability

All datasets generated for this study are included in the article/supplementary material.

## DATA AVAILABILITY STATEMENT

All datasets generated for this study are included in the article/supplementary material.

## ETHICS STATEMENT

The studies involving human participants were reviewed and approved by Zhangzhou Municipal Hospital Ethics Committee. Written informed consent for participation was not required for this study in accordance with the national legislation and the institutional requirements.

## AUTHOR CONTRIBUTIONS

WC and GH conceived the study and design, GH wrote the manuscript. SZ, DL, ZP, XZ, XL,RC, LZ collected the clinical data and following-up. TY, YW, WC supervised the project. All authors have read and approved the final manuscript.

